# Immunological Signatures During Type 2 Diabetes Development and Progression among Mexican Americans in Starr County, Texas

**DOI:** 10.1101/2025.06.23.25329883

**Authors:** Claudia C. Biguetti, Antonio Hernandes Chaves-Neto, Peter Elvin, Javier La Fontaine, Eric L. Brown, Craig L. Hanis, Walid D. Fakhouri

**Affiliations:** Department of Podiatric Medicine, Surgery and Biomechanics, School of Podiatric Medicine, The University of Texas Rio Grande Valley; Center for Craniofacial Research, Department of Diagnostic and Biomedical Sciences, The University of Texas Health Science Center School of Dentistry at Houston; Department of Epidemiology, Human Genetics and Environmental Sciences, School of Public Health, The University of Texas Health Science Center at Houston

**Keywords:** Diabetes Mellitus, Type 2, High Mobility Group Box 1 Protein, Mexican Americans, Insulin Resistance, Immunity

## Abstract

Previous studies implicate immune dysregulation in the metabolic changes accompanying obesity and type 2 diabetes. This study investigated the interplay between metabolic and immunological parameters during the progression of type 2 diabetes in an obese Mexican American cohort from Starr County, Texas. Individuals matched for age, gender, and BMI were stratified into five categories: diabetes-free, isolated impaired glucose tolerance, combined glucose impairment (fasting or post-load), type 2 diabetes without complications, and type 2 diabetes with lower extremity complications. Buffy samples were analyzed via Luminex Multiplex Assay for IL-4, IL-17A, MCP-1, and HMGB1. HMGB1 levels were significantly elevated in individuals with prediabetes and the combined glucose intolerance group compared to the diabetes-free group and those with diabetes. Elevated HMGB1 levels positively correlated with Homeostatic Model Assessment (HOMA) for insulin resistance (p = 0.03) and showed a moderate negative correlation with insulin sensitivity (p = 0.060). IL-17A levels were elevated in the diabetes group without complications compared to those with combined glucose impairment. The combined diabetes group exhibited the poorest glycemic control. In summary, HMGB1 is a potential early marker of insulin resistance and diabetes progression in Mexican Americans from an underserved community.

**Graphical Abstract:** 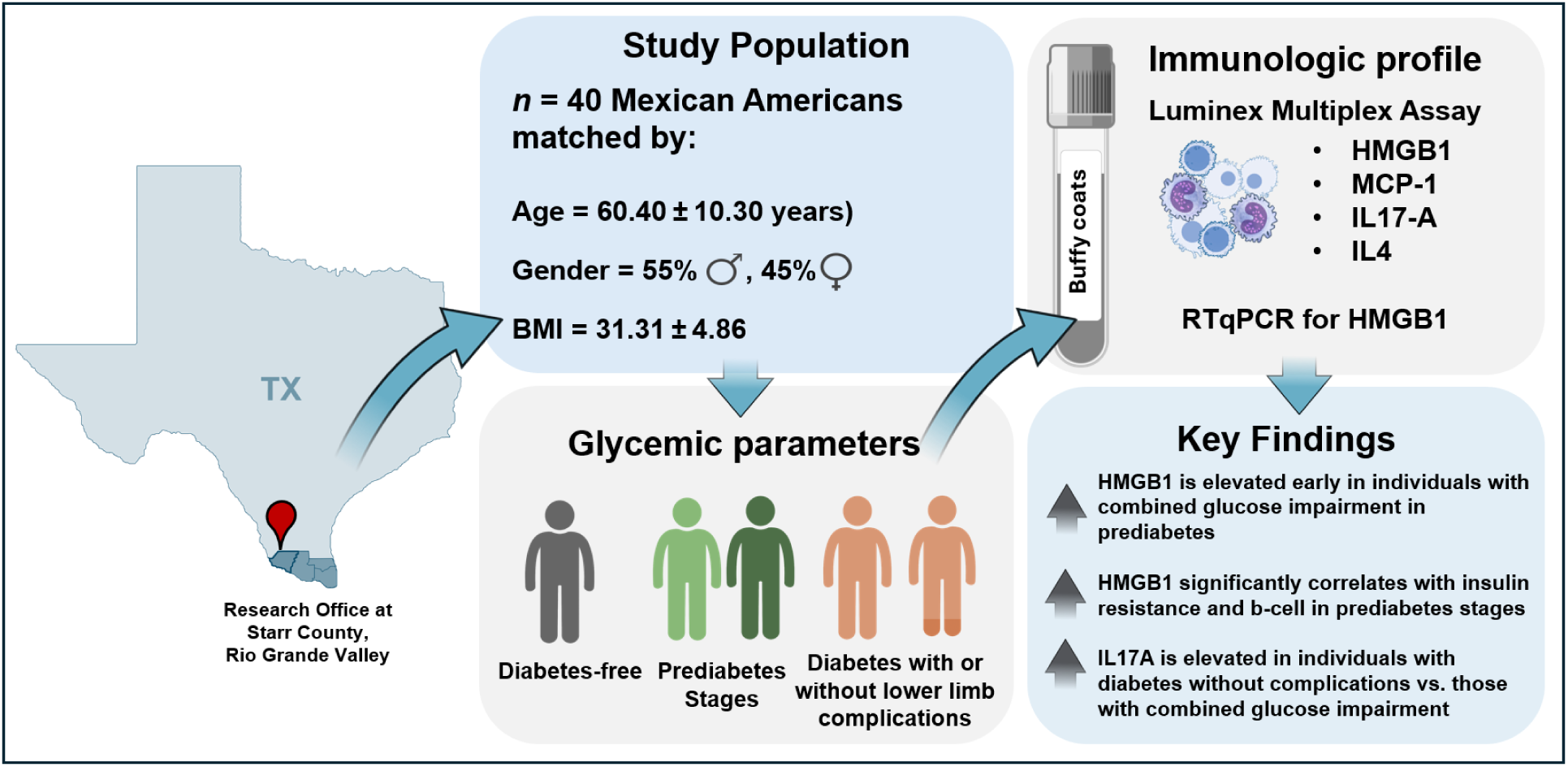

## Introduction

Type 2 diabetes is a growing global public health concern and leading contributor to mortality, reduced life expectancy, and increased healthcare burden^1,2^. In the United States alone, over 37 million adults are diagnosed with diabetes, with 90–95% of cases classified as type 2 diabetes^3^. In addition, 96 million American adults are considered to have prediabetes, with Hispanic or Latino population appearing at higher risk as compared to white non-Hispanic^4,5^. A pioneering study from 1983 in Starr County, a rural, medically underserved region on the border of Texas with Mexico, was among the first to document high rates of obesity and type 2 diabetes in this population ^6^. Between 2012 and 2022, diabetes prevalence in this region reached 43.95%, significantly higher than the national average of 26.73% ^7^. This disparity has been attributed to a combination of genetic, lifestyle, and socioeconomic factors, including higher rates of obesity, less access to healthcare, and cultural dietary patterns that may predispose this group to a higher risk of developing the disease ^6,8–15^. The convergence of these factors makes the Hispanic community along the US-Mexico border a critical area of focus for diabetes research, public health initiatives, and targeted interventions aimed at curbing the rising tide of this health crisis.

A growing body of evidence highlights the critical role of the immune system in the pathogenesis of obesity-related type 2 diabetes, with chronic low-grade inflammation being recognized as a hallmark of metabolic syndrome and insulin resistance^16,17^. In addition, studies have linked immunometabolic dysregulation in diabetes to alterations in the gut microbiome, which may impair mucosal immunity and promote systemic inflammation ^18^. Studies among Hispanics have demonstrated diabetes-associated shifts in both gut and salivary IgA microbiomes, with a potential immune dysregulation led by microbiota-mediated mechanisms in disease progression ^10,12^. Further studies confirmed gut microbiome changes linked to type 2 diabetes in this population, highlighting microbiota-immune interactions in this high-risk population ^19^. It is possible that circulating immunological proteins may serve as useful indicators of type 2 diabetes progression and complications. In Mexican Americans with type 2 diabetes, elevated serum levels of IL-6 and TNF-α have been associated with poorer glucose control^20^ and positively correlated with advanced complications such as diabetic retinopathy ^21^. However, successful investigations of immunological biomarkers and their correlation with progressive metabolic changes remain limited.

Beyond classical molecules implicated in metabolic inflammation and insulin resistance in obesity (e.g. MCP-1, IL6, IL17)^22^, High Mobility Group Box 1 (HMGB1) has recently gained attention due to its pro-inflammatory role under oxidative stress and chronic inflammatory states in different diseases ^23,24^. HMGB1 is a nuclear protein that, under homeostatic conditions, regulates chromatin structure and gene transcription^25,26^. However, upon cellular stress, injury, or metabolic dysregulation, HMGB1 is translocated from the nucleus to the cytoplasm and can be passively released from necrotic cells or actively secreted by immune cells into the extracellular space^23,26^. Its extracellular function is isoform-dependent, with most proinflammatory activity attributed to the disulfide isoform, which promotes inflammation by activating pattern recognition receptors such as TLR4 and RAGE, thereby amplifying innate immune responses^24,27^. More recently, HMGB1 has been implicated in the inflammatory milieu of diabetes complications, with elevated serum levels reported in patients with lower extremity complications, such as in peripheral artery disease^28^, and the highest HMGB1 concentrations observed in patients with type 2 diabetes and diabetic foot ulcers compared to those with type 2 diabetes but no foot ulcers^29^. However, its clinical relevance in high-risk Hispanic populations, particularly in relation to disease progression and diabetes complications, remains largely unexplored.

In this study, we estimated the correlation between immunological blood parameters and progression of type 2 diabetes in an obese Mexican American cohort from Starr County, Texas. Specifically, we analyzed demographic and clinical characteristics, biochemical markers of glucose metabolism, and levels of selected immunological markers in individuals across the type 2 diabetes spectrum. We hypothesized that diabetes progression in Mexican Americans is associated with a dysregulated immune response. To test this hypothesis, we employed a comparative analysis of matched subgroups for age, gender, and BMI, allowing us to isolate differences due to glycemic status rather than confounding demographic factors. A Luminex Multiplex Assay was conducted on buffy coat-derived samples to quantify key immune markers, accompanied by mRNA level of HMGB1. For HMGB1, we further performed correlation analyses to explore associations with HbA1c, as well as Homeostatic Model Assessment indices, including HOMA-IR (insulin resistance), HOMA-B (β-cell function), and HOMA-IS (insulin sensitivity).

## Results

The cohort for this study was comprised of 40 individuals, including 21 with no prior diagnosis of type 2 diabetes and 19 with a prior diagnosis. The 19 with a prior diagnosis were further stratified into those with no lower extremity complications and those with complications. Complications included a history of non-healing ulcers in the foot, ankle or lower leg with or without a history of amputation and hospitalization. The diabetes-free and diabetes groups were selected to be matched for age, gender, and BMI. The average age of participants was 60.40 ± 10.30 years, with a balanced gender distribution (55.00% male, 45.00% female). Overall mean BMI was 31.31 ± 4.86. Descriptive statistics are found in Table 1. In this cohort, 65% were born in Mexico with 35% born in the USA. In the diabetes group, 47.37% were from Mexico and 52.63% from the USA. For the diabetes-free group, a substantial majority of 80.95% were originally from Mexico, compared to 19.05% from the USA. Other variables in Table 1 include waist-to-hip ratio, fat-free mass using Segal et al.’s equations, usage of diabetes medications, current smoking status, and ethanol consumption. Waist-to-hip ratio was significantly higher in the diabetes group, but there were no differences between those with or without complications.

**Table 1.** Demographics and Clinical profile in individuals with or without diabetes from Starr County.

|  | No Diabetes | Prior diagnosis of diabetes | Diabetes without complications | Diabetes with complications |
| --- | --- | --- | --- | --- |
| **Age in years at the time of study (Mean ± SD)** | 62.19 ± 9.38 | 58.42 ± 11.15 | 59.57 ± 11.92 | 55.20 ± 8.93 |
| **Gender** |  |  |  |  |
| Male | 9 (21), 43% | 13 (19), 68% | 9 (14) | 4 (5) |
| Female | 12 (21), 57% | 6 (19), 32% | 5 (14) | 1 (5) |
| **Country of birth** |  |  |  |  |
| Mexico | 17 (21) 80.95% | 9 (19), 47.37% | 8 (14), 57.14% | 1 (5) 20% |
| USA | 4 (21) 19.05% | 10 (19), 52.63% | 6 (14) 42.86% | 4 (5) 80% |
| **BMI** | 31.03 ± 4.04 | 31.63 ± 5.73 | 31.60 ± 5.98 | 31.71 ± 5.58 |
| **Systolic Blood Pressure (Mean ± SD)** | 136.52 ± 19.56 | 146.53 ± 19.33 | 141.50 ± 16.91 | 160.60 ± 20.38 |
| **Diastolic Blood Pressure (Mean ± SD)** | 76.38 ± 9.51 | 80.53 ± 7.77 | 78.71 ± 8.10 | 85.60 ± 3.85 |
| **Waist to hip ratio** | 0.96 ± 0.07* | 1.01 ± 0.06* | 1.01 ± 0.07 | 1.01 ± 0.06 |
| **Segal et al. Equations**<br>**for FFM** | 47.52 ± 36.16 | 43.83 ± 52.33 | 48.47 ± 44.94 | 30.84 ± 73.98 |
| **Diabetes Medication**<br>**Usage** |  |  |  |  |
| *None* | 21 (21) | 2 (19), 10.53% | 2 (14) | 0 (5) |
| *Oral agent only* | 0 (21) | 11 (21), 52.38% | 10 (14) | 1 (5) |
| *Insulin only* | 0 (21) | 1 (21), 4.76% | 0 (14) | 1 (5) |
| *Oral agent AND Insulin* | 0 (21) | 5 (21), 23.81% | 2 (14) | 3 (5) |
| **Current Smoking Status** |  |  |  |  |
| *Currently smokes* | 2 (21), 9% | 5 (19), 26% | 3 (14) | 2 (5) |
| *Formerly smoked* | 5 (21), 24% | 3 (19), 16% | 2 (14) | 1 (5) |
| *Never smoked* | 14 (21), 67% | 11 (19), 58% | 9 (14) | 2 (5) |
| **Ethanol Consumption** | 4 (21), 19% | 7 (19), 37% | 5 (14) | 2 (5) |

Data is depicted as mean ± standard deviation (SD). Statistically significant differences ( $p < 0.05$ ) between groups are denoted by the asterisk symbol (\*).

The following analyses included detailed measurements of fasting and two-hour post-oral load blood glucose (mg/dL), fasting and two hour post-load plasma insulin (μU/mL), fasting and two hour post-load plasma c-peptide (ng/mL), and fasting plasma proinsulin (pM) and HOMA indices. These data were collected at the Research Office of the Starr County Health Studies in Rio Grande City, Texas (**Supplementary Table 1).**

Examining distributions within the group with no prior diagnosis of diabetes, we identified several individuals likely meeting the criteria for prediabetes. As a result, only 9 participants were classified as diabetes-free, while the remaining 12 were reclassified into various stages of prediabetes and subdivided (**Supplementary Table 2**). The stratification of diabetes by the presence or absence of lower extremity complications was maintained. According to this new stratification, 12 out of 21 individuals from the original diabetes-free group were classified with prediabetes and subdivided according to their stages: 2 with isolated impaired fasting glucose, 5 with isolated impaired glucose tolerance and 4 with combined impaired fasting glucose and impaired glucose tolerance. As only two individuals were identified with isolated impaired fasting glucose, they were included in the prediabetes group for combined analysis (Tables 2 and 3) but excluded from subgroup-specific analyses when considering immunological markers in buffy coats.

**Table 2.** Immunological markers levels in buffy coats from obese Mexican American individuals across different stages of type 2 diabetes progression.

| Group | HMGB1<br>pg/ml | IL-17A<br>pg/ml | IL-4<br>pg/ml | MCP-1<br>pg/ml |
| --- | --- | --- | --- | --- |
| Free of Diabetes | 1426.57±515.85 | 4.52±0.94 | 1.11±0.38 | 57.44±4.65 |
| isolated impaired Glucose Tolerance | 2124.40±56.62 | 4.77±0.66 | 1.20±0.18 | 65.39±20.97 |
| Combined Glucose Impairment | 4318.38±1552.19 | 3.76±1.27 | 1.00±0.57 | 49.27±21.85 |
| Diabetes without Complications | 2227.94±176.13 | 6.10±0.52 | 1.39±0.43 | 60.82±17.31 |
| Diabetes with Complications | 2209.03±1146.75 | 4.31±0.49 | 0.90±0.25 | 52.92±10.45 |
| ANOVA p-value | **0.0047\*** | **0.0151\*** | 0.4524 | 0.6689 |

*Data is expressed by Mean ± SD. Statistically significant differences ( $p < 0.05$ ) after ANOVA.*

Stratified analysis among diabetes free, prediabetes, and diabetes groups, identified several significant differences across these groups. HbA1c levels were significantly higher in the diabetes group compared to both the diabetes-free and prediabetes groups (p < 0.001) (**Supplementary Table 3**). Within the diabetes subgroups, participants with lower extremity complications exhibited significantly higher HbA1c values than those without complications (p = 0.0113), indicating poorer glycemic control. Additionally, fasting blood glucose levels were significantly elevated in the diabetes with complications subgroup compared to those without complications (p = 0.004). Although age, 2-hour post-load glucose levels, and years since diagnosis did not differ significantly among groups, metabolic and cardiovascular indicators showed additional trends (Supplemental Table 4). Waist-to-hip ratio was significantly higher in individuals with diabetes compared to those without (p = 0.0232) but had no significant relation to prediabetes. HDL cholesterol levels were significantly lower in the diabetes group relative to both those without and those with prediabetes (p = 0.0115). No statistically significant differences were found between the diabetes subgroups with or without complications for other cardiovascular parameters, including total cholesterol, LDL, triglycerides, and waist circumference.

To investigate whether these stages of metabolic dysfunction were accompanied by changes in immune profiles, immunological markers across the stratified groups were examined. Buffy coats from all 40 individuals were initially assessed for multiplex ELISA assays; however, after excluding values falling below detection limit, a subset of 22 individuals remained for statistical analysis on immunological markers.

Before analyzing cytokine expression, we re-evaluated insulin, glucose and HOMA values within this reduced cohort (**Figure 1**). In brief, HbA1c, fasting and 2-hour post-load glucose were significantly elevated in the diabetes groups compared to the diabetes free and prediabetes groups (p < 0.001, p = 0.00026, and p = 0.00221, respectively). Individuals with diabetes and complications exhibited the highest values for all three measures. C-peptide levels 2 hours post-glucose load also differed significantly among groups (p = 0.00112), with elevated levels in the combined glucose impairment group. HOMA was used to quantify insulin resistance (HOMA-IR), beta-cell function (HOMA-B), and insulin sensitivity (HOMA-S). The diabetes groups presented significantly lower beta-cell function compared to the prediabetes group. HOMA-IS was higher in those without diabetes compared to the diabetes groups, as expected.

**Figure 1.**
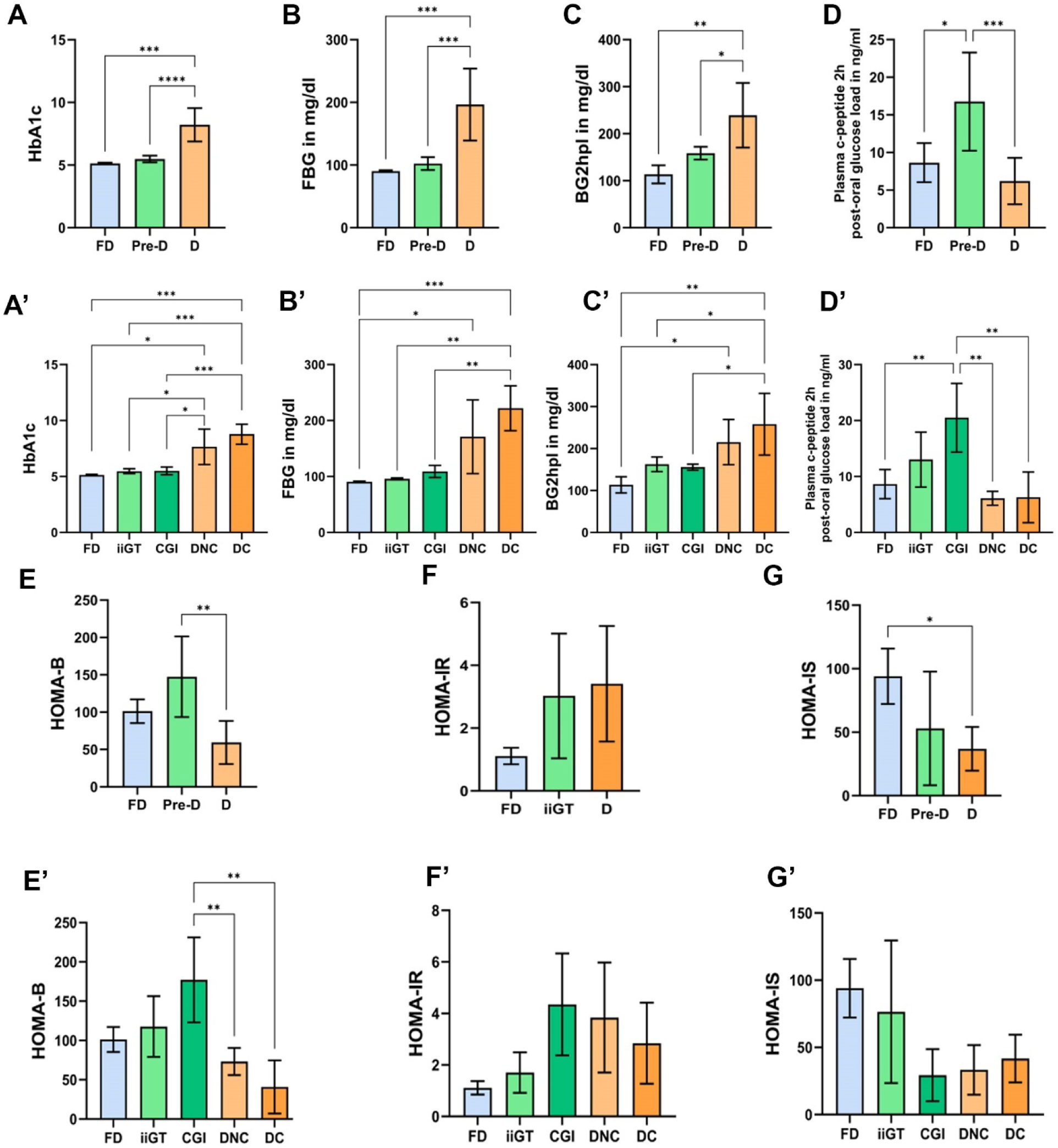
Comparison for glucose metabolism parameters in subgroups further considered for immunological analysis. Statistical analysis was conducted among individuals classified into different diabetes categories: Free of diabetes (FD), prediabetes (isolated impaired glucose tolerance (IGT) and combined glucose impairment (CGI)), and previously diagnosed diabetes (D). Within the diabetes group, individuals are distinguished between those without lower extremity complications (DNC) and those with such complications (DC). A–G summarize statistical analyses from the combined groups, while A’–G’ summarize the same results, but when stratified by subgroups. Data are expressed as Mean ± SD (Standard Deviation) and were analyzed using One-Way ANOVA followed by Tukey’s post hoc test. Significant differences, denoted by *, indicate p≤0.05.

The multiplex ELISA assay was performed for several immunological parameters using participants’ buffy coats. IL10, IL6, and TNF alpha presented lower levels in all groups, with most values being below the limit of detection (*data not shown*). The remaining marker results (HMGB1, IL-17A, MCP-1, and IL-4) are presented in **Table 2 and Figure 2**. Both IL-17A and HMGB1 showed statistically significant differences among the groups. IL-17A levels were elevated in the diabetes group (particularly in those with no complications) compared to the combined glucose impairment prediabetes group, although this significance was lost when the isolated impaired fasting glucose group was included. HMGB1 was significantly increased in prediabetes individuals compared to both the diabetes-free and diabetes groups. Interestingly, in the stratified analysis, HMGB1 levels were higher in the combined glucose impairment group compared to the isolated impaired glucose tolerance group, suggesting a potential association between elevated HMGB1 in buffy coats and specific stages of diabetes progression. No individuals with prediabetes were receiving any medications related to diabetes, whereas all patients in the diabetes group within this reduced cohort were under treatment (data not shown). IL-4 and MCP-1 levels did not show a significant difference across the groups. Interestingly, gene expression of HMGB1 did not show a difference amongst those free of diabetes compared to those with prediabetes or diabetes (Supplementary Figure 1). Since HMGB1 may be more indicative of type 2 diabetes progression, we focused on correlation analysis of this inflammatory marker.

**Figure 2.**
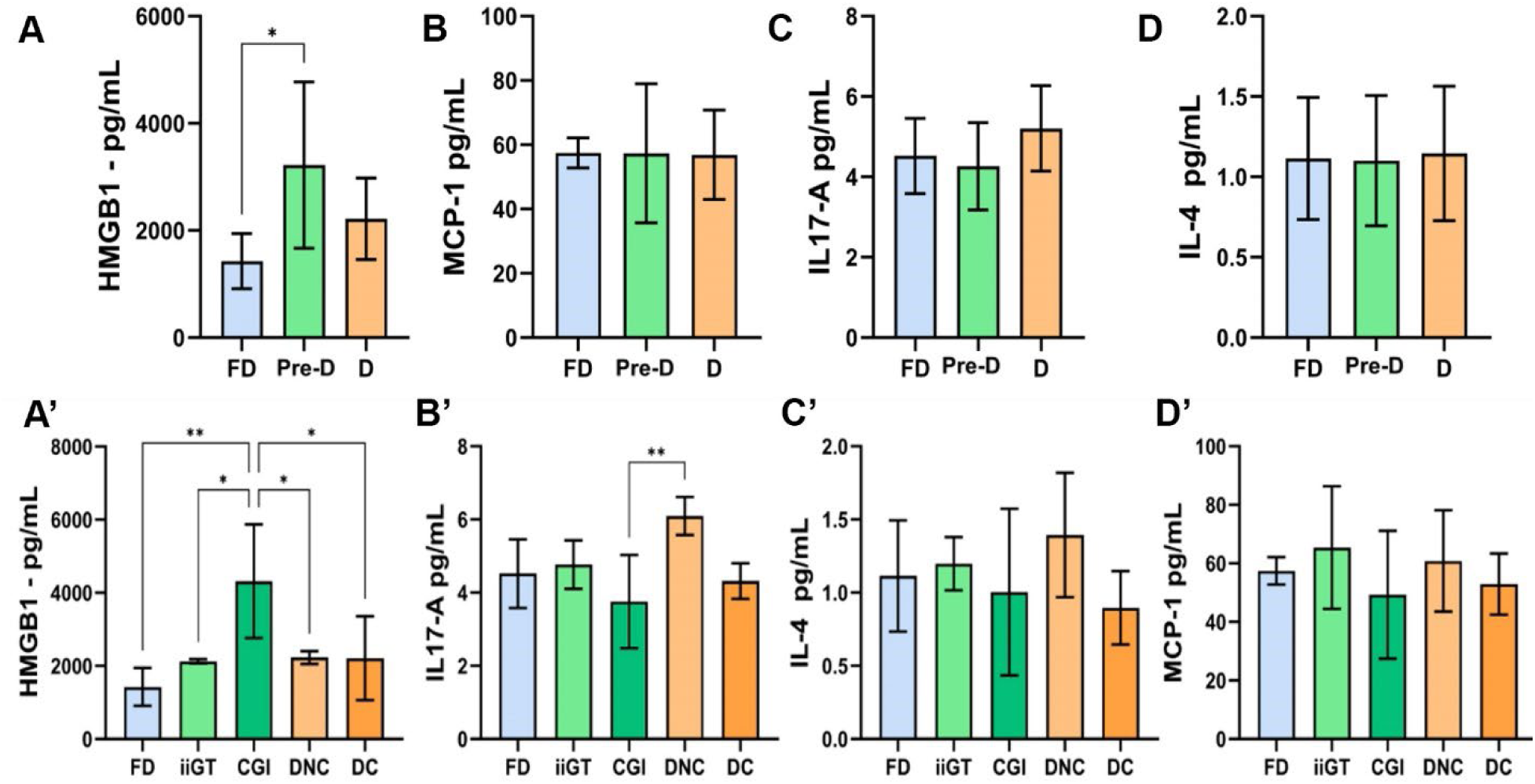
Comparison of immunological marker levels across diabetes progression groups. Statistical analysis was conducted among individuals classified as having no diabetes (FD), prediabetes (isolated impaired glucose tolerance (iiGT) or combined glucose intolerance (CGI)), or diabetes (with (DNC) or without (DC) complications). Data are presented as mean ± standard deviation (SD) and were analyzed using one-way ANOVA followed by Tukey’s post hoc test. Asterisks (*) indicate statistically significant differences (p ≤ 0.05). A–D present statistical analyses from combined groups, while A’–D’ present statistical analyses from the same data stratified by subgroups

To better elucidate the relationship between HMGB1 protein levels and the progression of type 2 diabetes, we conducted a series of correlation analyses with paired values across the diabetes and diabetes-free groups. These analyses examined the relationships between HMGB1 levels and HbA1c, HMGB1 and c-peptide levels (fasting and post-glucose challenge), and HMGB1 and the different HOMA parameters across the five stratified groups (Figure 3). The results indicate weak, non-significant correlation between HMGB1 and HbA1c (r = –0.04, p = 0.87) or fasting c-peptide (r = 0.17, p = 0.47) (Figure 3 A-B). However, a significant positive correlation was observed between HMGB1 and c-peptide levels at 120 minutes post-glucose load (r = 0.54, p = 0.02), indicating a potential link between HMGB1 and β-cell responsiveness during glucose challenge (Figure 3C). Additionally, HMGB1 showed a suggestive positive trend with HOMA-B (r = 0.396, p = 0.067), suggesting a possible association with β-cell function (Figure 3D). A negative correlation was observed between HMGB1 and HOMA-IS (r = –0.40, p = 0.06), indicating reduced insulin sensitivity with rising HMGB1 levels (Figure 3E). Importantly, HMGB1 levels were significantly and positively correlated with HOMA-IR (r = 0.45, p = 0.03), reinforcing its potential role as an immunometabolic marker linked to insulin resistance (Figure 3F). Furthermore, individuals in the prediabetes group of combined glucose impairment appeared to contribute most strongly to the observed correlations, exhibiting elevated HMGB1 levels coupled with reduced insulin sensitivity (low HOMA-IS), increased insulin resistance (high HOMA-IR), and compensatory β-cell activity (elevated HOMA-B). It is worth noting that, at the time of sample collection, these individuals were classified as not having diabetes due to their declaring no history of diabetes and no usage of diabetes-related medications. These results suggest the potential of HMGB1 as an early biomarker of immunometabolic dysfunction during this evolving pre-diabetes phase. Finally, in the diabetes without complications group included in the immunological analysis, one participant was not taking any diabetes-related medications, while the remaining participants were on oral agents only. In contrast, participants in the diabetes with complications group were either on both oral and insulin therapy or on one of these treatments alone.

**Figure 3.**
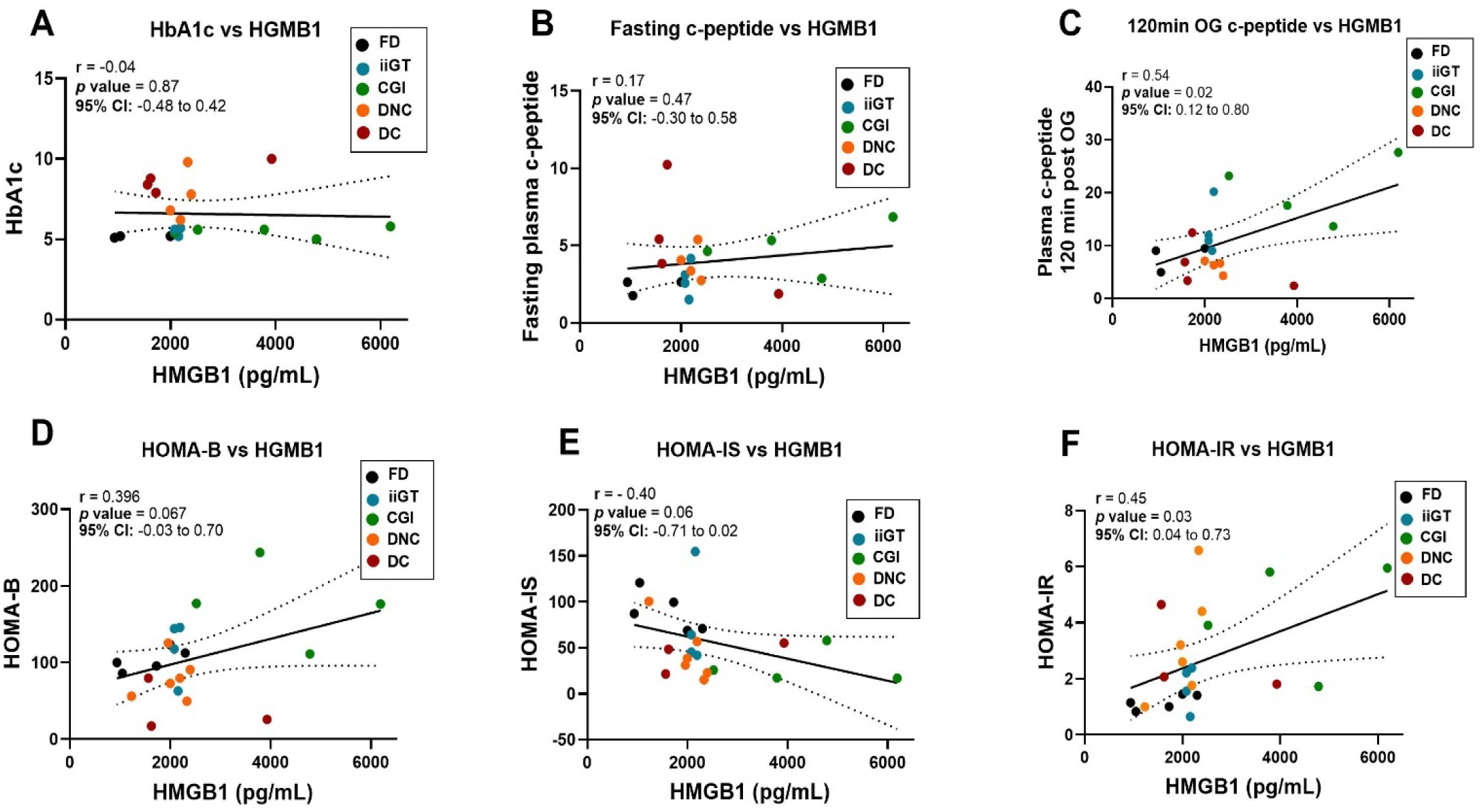
Pearson correlation analysis between HMGB1 concentrations in buffy coats and various indicators of glycemic control and insulin function in plasma. Panels A–C show correlations between HMGB1 and HbA1c, fasting plasma c-peptide, and 120-minute post-glucose load c-peptide, respectively (n = 19). Panels D–F display correlations between HMGB1 and HOMA-B (β-cell function), HOMA-IS (insulin sensitivity), and HOMA-IR (insulin resistance), respectively (n = 22). Each data point represents an individual classified as free of diabetes (FD), or having prediabetes (isolated impaired glucose tolerance (iiGT) or combined glucose impairment (CGI)), or Diabetes (with (DC) or without (DNC) complications). Solid lines indicate linear regression fit, and dotted lines represent the 95% confidence interval. Statistical significance was defined as p ≤ 0.05.

## Discussion

This study provided a unique opportunity to explore immunological parameters in progression of diabetes in an obese Mexican American population from the Texas-Mexico border, an underserved and underinsured population disproportionately affected by metabolic disease^7,15,30^. Our findings build on existing evidence linking chronic low-grade inflammation to metabolic dysfunction^20,21^ and underscore the potential of HMGB1, an alarmin associated with innate immune activation, as a potential early immunometabolic marker of insulin resistance in high-risk individuals^22,31–36^.

Upon examining the standard deviation and distribution of values in the diabetes-free group (**Supplemental Figure 1**), we identified certain individuals potentially fell into the pre-diabetes category. It has been considered that impaired glucose tolerance and impaired fasting glycemia represent intermediate stages in the transition from normal glycemia to type 2 diabetes^37,38^. Thus, in order to increase the specificity in our statistical analysis for both diabetes-free and diabetes groups, participants were further stratified into subcategories (**Table 2)**, consistent with previous studies^39^ and the latest updates of Standards of Care in Diabetes from the American Diabetes Association^37^. Consequently, our cohort stratification was based on previous clinical diagnoses of type 2 diabetes, oral glucose tolerance tests, and other biochemical/metabolic parameters. The group with diabetes was further stratified according to the presence or absence of lower extremity complications. The inclusion of lower extremity complications served as a critical indicator of advanced disease, even though no significant differences were observed in age (59.57 ± 11.92 vs. 55.2 ± 8.93 years) or duration of diabetes diagnosis (15.20 ± 7.53 vs. 12.93 ± 8.35 years) between participants without complications and those with complications (**Supplementary Table 3**). Lower extremity complications, such as diabetic foot ulcers and amputations, are among the most severe consequences of poorly managed type 2 diabetes and are associated with reduced mobility, diminished quality of life, and substantially decreased life expectancy^40^. Indeed, the presence of lower extremity complications with diabetes correlated with the highest mean values for fasting blood glucose and 2-hour postprandial glucose, suggesting that these complications are associated with poorer glycemic control. Interestingly, the mean HbA1c levels were higher in the diabetes with no complications group than in those with complications, which may reflect differences in treatment strategies, as most individuals in the complications group were receiving both oral and insulin therapy. This apparent discrepancy could also be influenced by greater glycemic variability in the complications group, resulting in elevated fasting and postprandial glucose despite a lower average HbA1c. In addition, the limited sample size of this pilot study may have contributed to variability in group means.

Sample stratification revealed a subset of individuals originally classified as diabetes-free displaying metabolic profiles consistent with prediabetes stages evolving to type 2 diabetes. According to the literature, HOMA-IR values greater than 2.5 are often considered indicative of insulin resistance, while HOMA-B values provide insight into beta cell function, with higher values indicating better beta cell function^39^. In addition, HOMA-IS values are useful to inform how effectively insulin is working to lower glycemia^39^. In this context, lower HOMA-IS values suggest reduced insulin sensitivity, which can be a characteristic of conditions such as insulin resistance during prediabetes stages and type 2 diabetes. With the results from the comparison amongst several biochemical parameters across the different groups, we infer that prediabetes groups have a compensatory increase in insulin secretion compared to the diabetes-free group. Within the prediabetes group, individuals who met the criteria for both impaired fasting glucose and elevated 2-hour post-load glucose (the combined glucose impairment group) exhibited elevated HOMA-B values, indicating a compensatory increase in β-cell activity in response to rising insulin resistance during the intermediate stages of diabetes progression.

Among all immunological markers tested in buffy coats, IL-17A levels were significantly increased in the group with diabetes without complications compared to the group with combined glucose impairment. The predominant source of IL-17A is the Th17 cell subset, a distinct lineage of CD4+ lymphocytes, which, in addition to IL-17A, secretes a variety of other pro-inflammatory cytokines involved in tissue inflammation and immune dysregulation^41^. This cytokine plays a role in chronic inflammation associated to diabetes complications^42–45^, including impaired wound healing in infected foot ulcers, further associated with increased T cell infiltration in individuals with advanced dysglycemia ^45^. Another previous study with obese Mexican American individuals from Starr County demonstrated that individuals with prediabetes and diabetes (collectively characterized as dysglycemia groups) revealed significantly higher Th17:Treg ratios in dysglycemia states compared to normoglycemia^10^. In that study, IL-17A-producing Th17 cells were quantified based on demethylated IL17A gene regions, allowing precise estimation of circulating Th17 cell populations from buffy coat samples across individuals with or without dysglycemia^10^. In individuals with chronic disease states like diabetes, increased Th17:Treg ratios correlated with changes in the gut microbiome, suggesting that Th17-driven inflammation may influence microbial composition^10^. Together, previous studies provide strong evidence that Th17 cell expansion (producing IL-17A) may contribute to systemic immunometabolic dysregulation in individuals with diabetes, associated with dysbiosis and disease progression^10,42,44,45^.

Of note, HMGB1 showed the most pronounced differences across different groups. Individuals categorized as having prediabetes based on both fasting and post-load glucose showed immunometabolic characteristics in HMGB1 levels most strongly associated with disease progression. Of note, the prediabetes stratification was established within this study based on metabolic profiling, and participants in this diabetes-free group had not yet received a formal diabetes diagnosis at the time of sample collection. As these individuals reported not being under pharmacological management, the observed elevation in HMGB1 may reflect immune alterations associated with early stages of disease progression. This suggests that HMGB1 may be elevated prior to the clinical diagnosis of diabetes, supporting its role as a potential early biomarker for disease risk^36^. Previous studies have confirmed HMGB1’s association with obesity ^32,33^, insulin resistance and hyperglycemia ^34^, inflammation^33,35^, and metabolic syndrome^36^, and increased cardiovascular disease risk ^41^. Our findings extend these observations by showing that HMGB1 levels in buffy coats also correlate with dynamic changes in metabolic function among Mexican Americans at various stages of type 2 diabetes. It is important to emphasize that gene expression analysis of HMGB1 did not reveal significant differences amongst the groups, despite a similar trend, suggesting that protein-level changes in buffy coats may serve as a more sensitive indicator of metabolic-immune shifts than transcriptional changes for this marker. Indeed, HMGB1 is a constitutively expressed nuclear protein involved in DNA organization, but its pro-inflammatory role emerges when it translocates from the nucleus to the cytoplasm, undergoes redox modifications, and is either actively secreted by immune cells or passively released from damaged cells into the extracellular space^23,24,26,42^.

Correlations between HMGB1 and metabolic markers further reinforce its relevance. A strong positive correlation between HMGB1 and 2-hour c-peptide levels links immune activation and late-phase insulin secretion. Additionally, the observed correlation between HMGB1 and HOMA-IR (r = 0.45, p = 0.03) and the negative trend with HOMA-IS (r = - 0.40, p = 0.06) imply that increased HMGB1 may signal declining insulin sensitivity and rising insulin resistance. While correlations with HbA1c and fasting c-peptide were not significant, the stronger associations with dynamic measures (post-load c-peptide and insulin indices) may reflect the greater sensitivity of these markers to early metabolic dysregulation. Our findings align with emerging literature on immunometabolism and the role of damage-associated molecular patterns such as HMGB1 in bridging metabolic stress and immune activation in obesity^31^. Inflammation-mediated HMGB1 release has been shown to perpetuate innate immune signaling via TLR4 and RAGE, amplifying insulin resistance in adipose tissue and promoting vascular complications^28,29,35,36,41^. The current study supports this framework, highlighting HMGB1’s potential as a dual marker of early immune perturbation and metabolic impairment. Taken together, these comparisons highlighted HMGB1 as a potential early indicator of immunometabolic dysregulation in high-risk individuals. The identification of accessible immune markers such as HMGB1 may facilitate earlier intervention and tailored prevention strategies in this underserved group.

In summary, this study highlights the potential value of buffy coat–derived immunological markers for detecting early immunometabolic alterations associated with type 2 diabetes progression. However, several limitations must be considered. The cross-sectional design limits causal inference, and the relatively small sample size constrained subgroup analyses, particularly regarding sex- and treatment-specific effects. While buffy coat samples offer practical advantages in clinical and translational settings, future studies incorporating plasma cytokine profiling, immune cell phenotyping, and longitudinal follow-up will be necessary to validate and expand upon these findings. Collectively, these results provide a foundation for further research aimed at improving early detection strategies and developing targeted interventions to reduce the burden of type 2 diabetes in high-risk Mexican American populations.

## Methods

### Study population

Data and samples from a total of 40 participants matched for age, gender and BMI were provided by the ongoing “Starr County Health Studies” (1981–present), conducted on the Texas-Mexico border. Participant information, samples, and medical records were obtained under written informed consent from each participant in accordance with the Institutional Review Board of the University of Texas Health Science Center prior to the initial submission of this pilot project and under the IRB: HSC-SPH-06-0225. In addition to the previous IRB, the Institutional Review Board of the University of Texas Rio Grande Valley approved the study (UTRGV-IRB-22-0273). All methods were performed following the relevant guidelines and regulations. All data were de-identified to ensure that no confidential information was included in the processed dataset. In brief, participants are enrolled and evaluated through interviews and medical exams conducted by a trained bilingual field team at the Rio Grande City field office, established in 1981. For the present research, a new exempt IRB was approved at the University of Texas Rio Grande Valley (UTRGV) under protocol IRB-22-0273. Inclusion criteria were: age > 40 years; no reported cancer, diabetes, or cardiovascular disease at the time of cohort entry. Exclusion criteria included participants with conditions that cause immunocompromise, such as HIV infection or current use of immunosuppressive medication.

### Buffy coat preparation for ELISA Multiplex

Buffy coat samples used in this study were obtained during a prior cohort assessment conducted at the Starr County Research Office. Blood samples were collected by trained personnel via venipuncture, placed on ice and processed on-site within 30 minutes to isolate buffy coats, which were subsequently preserved at –80°C until further analysis. The samples were later shipped on dry ice to the University of Texas Health Science Center at Houston School of Dentistry for protein extraction and quality assessment. Total protein was extracted using an ultrasonicator at 35% amplitude and 4 joules power, where the samples were kept on ice all the time, then centrifuged at 4°C for 15 minutes to remove non-soluble substances. Protein concentration was quantified using the Bradford Assay and a Qubit fluorometer to ensure accuracy and quantity across samples. Immunological proteins were quantified using the Luminex Multiplex Assay, conducted at the ORION Core Laboratory of MD Anderson Cancer Research Center. The assay employed the MILLIPLEX® Human Cytokine/Chemokine/Growth Factor Panel (HCYTA-60K, Millipore Sigma), which included IL-4, IL-6, IL-10, IL-17A, TNF-α, and MCP-1. Another plate from the MILLIPLEX® Human Cytokine was customized for the HMGB1 protein. After data acquisition, samples with biomarker concentrations falling outside the assay’s detection range were excluded from analysis. The final sample size for ELISA multiplex analysis was 22 individuals, depending on the specific marker evaluated.

### RTqPCR for HMGB1

Total RNA was extracted from blood samples using the TRIzol solution, followed by acidic phenol/chloroform for purification. The RNA was precipitated using cold isopropanol, tRNA, and 30 mM sodium acetate. The mRNA expression of *HMGB1* was measured in 22 samples using five technical replicates per sample. We used the following qPCR primers to quantify the mRNA level for *HMGB1.*F: ATCCCAATGCACCCAAGAGG and *HMGB1.*R: GACAGGCCAGGATGTTCTCC using SYBR-Green Gold Taq-Polymerase. We also use primers for GAPDH F: AATGGGCAGCCGTTAGGAAA and GAPDH.R: AGGAAAAGCATCACCCGGAG, as a housekeeping gene for normalization.

### Statistical Analysis

All statistical analyses were performed using GraphPad Prism version 10 (GraphPad Software, San Diego, CA). Descriptive statistics for continuous variables are presented as mean ± standard deviation (SD). Data normality was assessed using the Shapiro-Wilk test. For comparisons across three or more groups, one-way ANOVA was used, followed by Tukey’s post hoc test for multiple comparisons when data met parametric assumptions. For non-normally distributed variables or unequal variances, the Kruskal–Wallis test with Dunn’s multiple comparisons was applied. Comparisons between two independent groups (e.g., diabetes-free vs. diabetes) were conducted using unpaired two-tailed t-tests for normally distributed data, or Mann-Whitney U tests for non-parametric distributions. Categorical variables such as medication use or origin (Mexico vs USA) were compared using Chi-square tests or Fisher’s exact test, as appropriate. Pearson correlation analyses were used to examine associations between HMGB1 levels and metabolic indices (e.g., HOMA-IR, HOMA-B, HOMA-IS, HbA1c), with results expressed as correlation coefficients (r), 95% confidence intervals, and p-values. Correlation strength was interpreted as follows: r < 0.3 = weak, 0.3–0.5 = moderate, and > 0.5 = strong. Spearman’s correlation was applied when the data did not meet parametric assumptions. Outlier values beyond the detection range of the multiplex ELISA assay were excluded from analyses, and sample sizes for each comparison are reported in the figure and table legends. Statistical significance was defined as p ≤ 0.05, and values between p = 0.05 and 0.10 were interpreted as trends. Analyses were conducted across stratified groups: diabetes-free), prediabetes (isolated impaired glucose tolerance or combined glycose impairment), and diabetes (with or without complications), to capture immunometabolic variation across progressive disease states.

## Data Availability

All data produced in the present study are available upon reasonable request to the authors

## Data availability

The datasets used and/or analyzed during the current study are available from the primary corresponding author on reasonable request.

## Ethics Statement

All studies involving human participants were reviewed and approved by the Institutional Review Board of the University of Texas Health Science Center (HSC-SPH-06-0225) and the University of Texas Rio Grande Valley (UTRGV-IRB-22-0273). All participants provided written informed consent prior to their inclusion in the study.

## Funding

This work was supported by National Institutes of Health grants DK073541 and HL102830 to CLH and AI085014 to ELB and CLH, the UTSD Excellence in Dental Diagnostic Clinical Research Award to WDF, and NIH-NCATS 5UL1TR003167-04 to CCB, WDF, and CLH.

## Author Contributions Statement

C.C.B., W.D.F., E.L.B., and C.L.H. conceptualized the project, secured funding, provided resources and oversaw project administration and management. C.C.B. and P.E. conducted data analysis and prepared the tables and figures. W.D.F. and A.H.C.N. carried out the molecular analyses. C.L.H. and E.L.B. supervised all phases of data and sample collection, as well as project administration, from the Starr County Research Office. J.L.F. provided clinical insight and feedback on project conceptualization, data analysis, and manuscript drafting. All authors reviewed and approved the final manuscript.

## Acknowledgments

We extend our sincere appreciation to the dedicated staff at the Starr County Health Studies Field Office, whose continued commitment and expertise have been essential to the success of this research. We are also deeply grateful to all study participants for their valuable contributions and for entrusting their time and information in support of this work.

## Supplementary Information

**Supplementary Table 1.** Glucose Metabolism and Insulin-Related Parameters Between ND and D Groups.

|  | No Diabetes | Prior Diagnosis of Diabetes | *p-value* |
| --- | --- | --- | --- |
| Fasting blood glucose in mg/dl | 99.33 ± 8.60 | 163.89 ± 57.11 | 0.00017* |
| Blood glucose 120 minutes post-oral glucose load in mg/dl | 127.90 ± 31.94 | 209.83 ± 51.21 | 0.00000* |
| Fasting Plasma insulin in uU/ml | 16.06 ± 10.07 | 17.40 ± 10.58 | 0.69054 |
| Plasma insulin 120 minutes post-oral glucose load in uU/ml | 98.96 ± 87.70 | 43.02 ± 46.00 | 0.01616* |
| Fasting Plasma c-peptide in ng/ml | 3.59 ± 1.52 | 3.86 ± 1.33 | 0.54863 |
| Plasma c-peptide 120 minutes post-oral glucose load in ng/ml | 12.95 ± 6.48 | 6.62 ± 4.09 | 0.00076* |
| Fasting Plasma proinsulin in pM | 14.25 ± 12.15 | 29.16 ± 18.95 | 0.00770* |
| HOMA Model Steady State Beta Cell function | 135.11 ± 43.37 | 75.87 ± 46.78 | 0.00025* |
| HOMA Model Steady State Insulin Sensitivity | 57.84 ± 34.70 | 43.07 ± 22.59 | 0.1196 |
| HOMA Model insulin resistance | 2.41 ± 1.49 | 3.04 ± 1.74 | 0.23407 |

Data is depicted as mean ± standard deviation (SD). Statistically significant differences ( $p < 0.05$ ) between groups are denoted by the asterisk symbol (\*).

**Supplementary Table 2.**
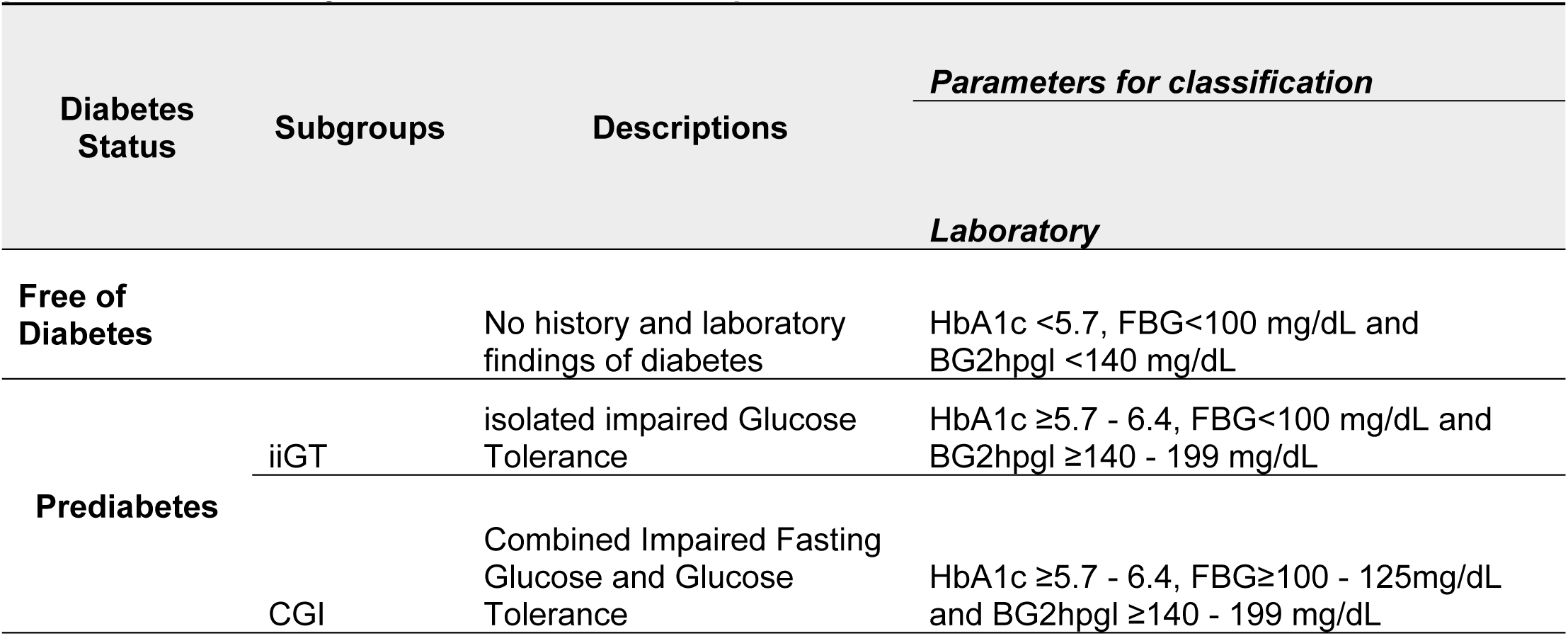
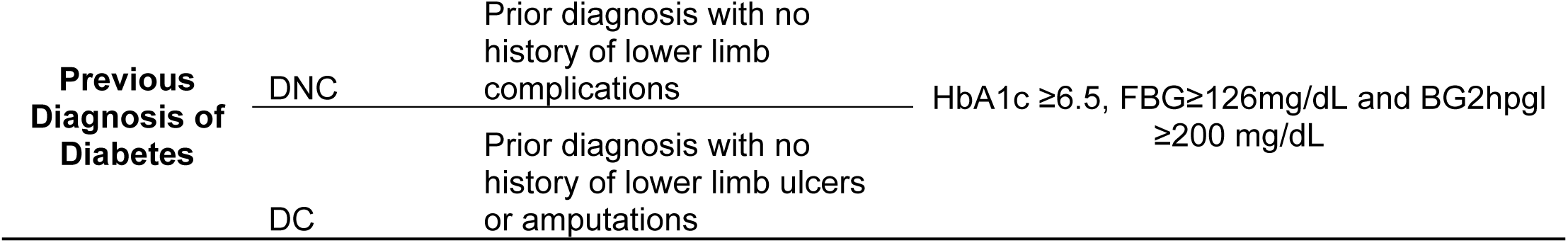
Criteria for subgroup stratification according to glycemic parameter and history of lower extremities complications.

**Supplementary Table 3.** Diabetes related parameters across Diabetes Progression.

| Group | Age (years) | HbA1C* | FBG (mg/dl) | BG2hpgl (mg/dl) | Years of Diabetes Dx |
| --- | --- | --- | --- | --- | --- |
| **Free of Diabetes** | 62.11±10.03 | 5.39±0.23* | 93.89±3.92 | 102.89±20.07 | N/A |
| **Prediabetes** | 62.25 ± 9.31 | 5.49±0.25<sup>#</sup> | 103.42 ± 8.99 | 146.67 ± 25.81 | N/A |
| **Diabetes** | 58.42±11.15 | 7.73±1.37*<sup>#</sup> | 167.89±58.18 | 217.74±60.53 | N/A |
| **p value** | *p* > 0.05 | < 0.001 | *p* > 0.05 | *p* > 0.05 | - |
| **DNC** | 59.57 ± 11.92 | 7.34 ± 1.34<sup>a</sup> | 145.64 ± 46.75<sup>a</sup> | 199.79 ± 48.46 | 15.20 ± 7.53 |
| **DC** | 55.2 ± 8.93 | 8.82 ± 0.78<sup>b</sup> | 230.20 ± 39.29<sup>a</sup> | 268.00 ± 67.59 | 12.93 ± 8.35 |
| **p value** | 0.413 | 0.0113 | 0.004 | 0.0872 | 0.5897 |

\*HbA1C was measured for 7 individuals out of 9 'Free of Diabetes'
Data is expressed by Mean ± SD. Statistically significant differences ( $p < 0.05$ ) between groups (Free Diabetes, Pre-diabetes and Diabetes) are denoted by the asterisk symbol (\*). Statistical differences between DNC and DC, if applicable, are denoted by different lowercase letters. FBG: Fasting Blood Glucose; BG2hpgl: Blood Glucose 2 Hours Post-Oral Glucose Load; DC: Diabetes with lower limb complications; DNC: Diabetes without lower limb complications.

**Supplementary Table 4.**
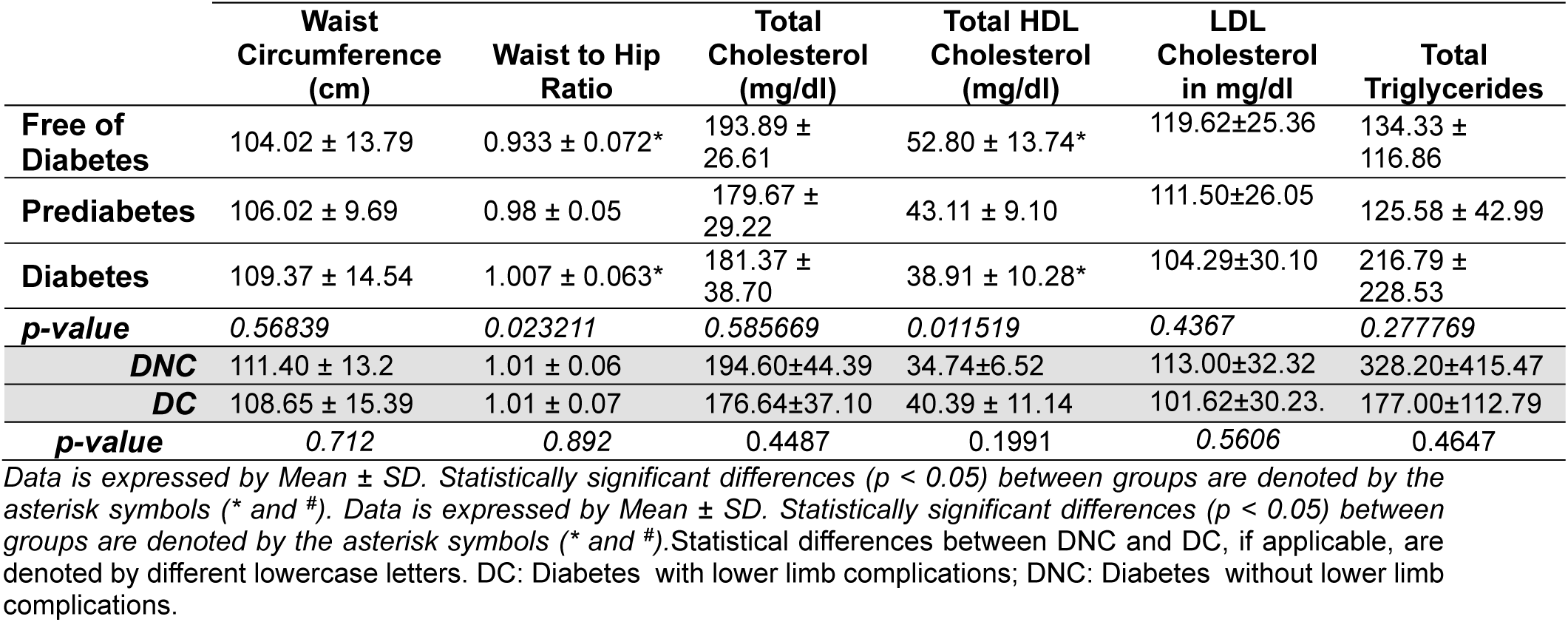
Cardiovascular Health Associated Parameters across Diabetes Progression.

**Supplementary Figure 1.**
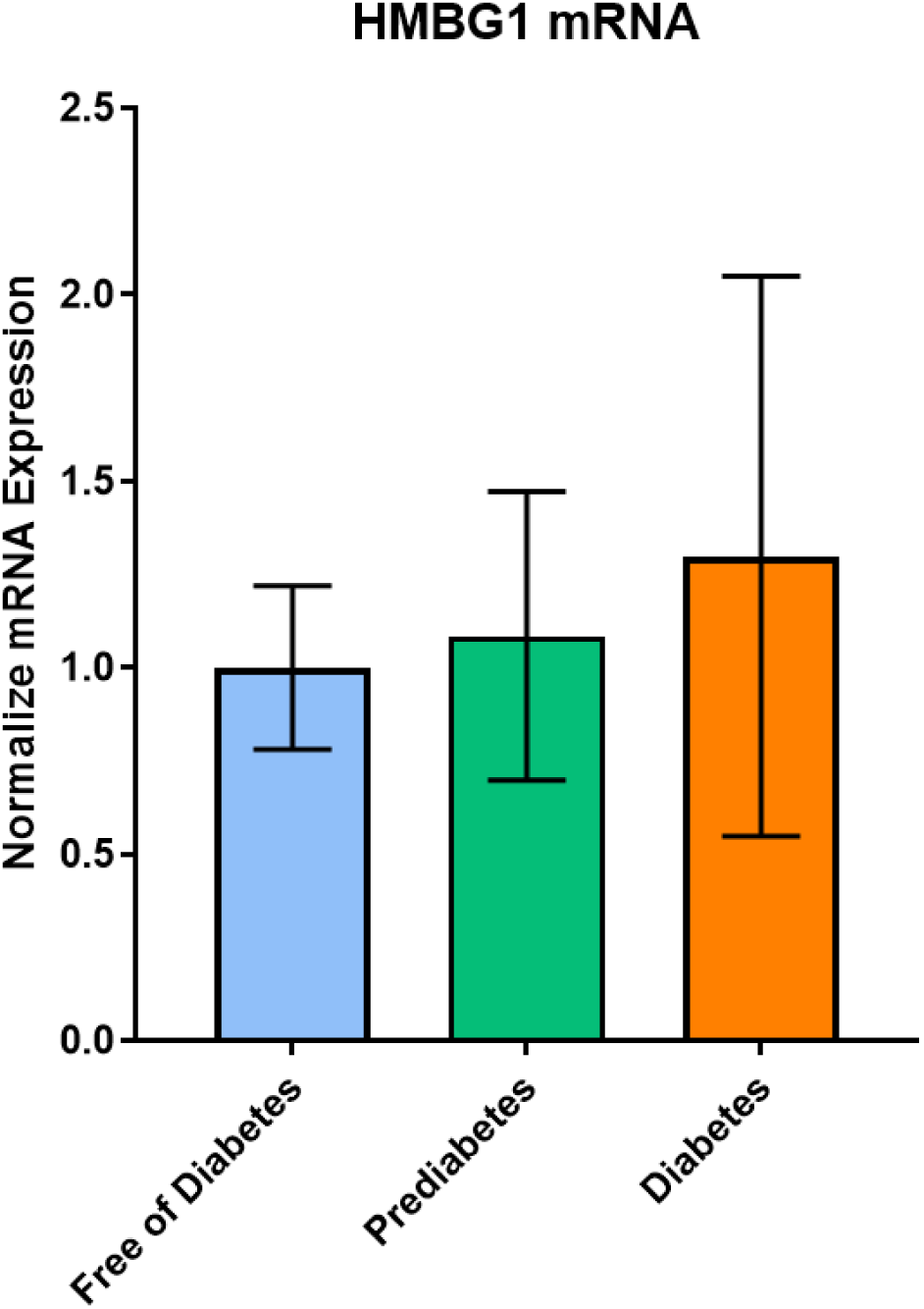
RTqPCR for 8 controls and 8 patients from each subgroup (FD, Pre-D and D). Statistical analysis employing One Way ANOVA followed by Tukey for Multiple Comparison. Abbreviations: FD = Free of Diabetes; Pre-D = Pre-Diabetes; D = Individuals with prior diagnosis of diabetes, including those with and without lower extremities complications.

## Notes

### Competing Interest Statement

The authors have declared no competing interest.

